# Immunogenicity of NVX-CoV2373 in PREVENT-19: A Phase 3, Randomized, Placebo-Controlled Trial in Adults in the United States and Mexico

**DOI:** 10.1101/2023.05.08.23289670

**Authors:** Germán Áñez, Karen L. Kotloff, Cynthia L. Gay, Joy Nelson, Haoua Dunbar, Shane Cloney-Clark, Alice McGarry, Wayne Woo, Iksung Cho, Joyce S. Plested, Gregory M. Glenn, Lisa M. Dunkle, the 2019nCoV-301 study group

## Abstract

**Background:** NVX-CoV2373, an adjuvanted, recombinant SARS-CoV-2 spike (rS) protein vaccine, consistently demonstrated protective efficacy against COVID-19 in clinical trials and has received regulatory authorizations or approvals worldwide.

**Methods:** PREVENT-19 (NCT04611802) is a phase 3, randomized, observer-blinded, placebo-controlled trial evaluating safety, immunogenicity, and efficacy of NVX-CoV2373 in ≈30 000 participants ≥18 years in the United States and Mexico. Vaccine humoral immune response (ie, serum immunoglobulin [IgG] antibodies, hACE2 receptor binding inhibition antibodies, and neutralizing antibodies to SARS-CoV-2) (ancestral strain) was assessed in 1200 participants randomly selected and equally divided between participants 18–64 and ≥65 years.

**Results:** In the per protocol analysis, NVX-CoV2373 induced vigorous serum antibody responses among the 1063 analyzed participants who were SARS-CoV-2 seronegative at baseline, received both doses of study treatment, and had serology results available 2 weeks after dose 2. Geometric mean (GM) responses in both younger and older adults were higher among recipients of vaccine versus placebo for IgG (64 259 vs 121 and 37 750 vs 133 ELISA units, respectively), hACE2 receptor binding inhibition GM titers (GMTs) (222 vs 5 and 136 vs 5, respectively), and neutralizing antibody GMTs (1303 vs 11 and 900 vs 11, respectively). Humoral responses were 30–40% lower in participants ≥65 years or HIV-positive; however, seroconversion rates were high and comparable between the age cohorts, regardless of HIV serostatus.

**Conclusions:** NVX-CoV2373 elicited robust humoral immune responses against ancestral SARS-CoV-2 virus 2 weeks following the second vaccination in adult PREVENT-19 participants, consistent with previously reported high vaccine efficacy.

PREVENT-19 ClinicalTrials.gov number, NCT04611802

---

Severe acute respiratory syndrome coronavirus 2 (SARS-CoV-2) spike (S) protein-based vaccines have been developed at an extraordinary pace since the declaration of the coronavirus disease 2019 (COVID-19) pandemic in early 2020 [1]. Over 35 vaccines have been approved by at least one country and 13 have received Emergency Use Listing (EUL) by the World Health Organization (WHO) based on assessment of safety, efficacy, and quality [2]. Vaccines using four platforms have been granted EUL: messenger RNA [3, 4], adenoviral vectors [5, 6], inactivated virus [7–11], and recombinant protein subunits [12, 13]. These vaccines have proven to be highly efficacious/effective for the prevention of severe disease in clinical trials [3–13].

Protection against less severe symptomatic infection is also robust (generally exceeding 50% and often ≥90%), albeit more variable depending on factors, such as vaccine components, study locations, and circulating viral variants [14].

NVX-CoV2373 is a SARS-CoV-2 recombinant S-protein vaccine adjuvanted with saponin-based adjuvant (Matrix-M™), which has been demonstrated to be safe and efficacious in clinical trials involving over 49 000 participants from Mexico, South Africa, the United Kingdom, and the United States [15–17]. These data have supported authorization in multiple regions and EUL by the WHO [18].

PREVENT-19 (<u>PRE</u>-fusion protein subunit <u>V</u>accine <u>E</u>fficacy <u>N</u>ovavax <u>T</u>rial COVID<u>-19</u>, NCT04611802), a large pivotal phase 3, randomized, placebo-controlled, multicenter clinical trial of NVX-CoV2373, demonstrated 90% vaccine efficacy (VE) against mild, moderate, or severe COVID-19 (primary endpoint) and 100% VE against moderate-to-severe disease caused by a variety of circulating SARS-CoV-2 variants among ≈30 000 adults ≥18 years of age in the United States and Mexico [17]. We report here the humoral immune responses elicited by NVX-CoV2373 in the demographically diverse participants in the PREVENT-19 study.

## METHODS

### Study Population

A total of 29 945 adults participating in PREVENT-19 were randomized in a 2:1 ratio to receive 2 intramuscular injections of either NVX-CoV2373 or placebo 21 days apart [17].

Immunogenicity testing was performed on a subset of ≈1200 randomly selected participants (regardless of previous SARS-CoV-2 infection, indicated by anti-nucleocapsid antibody or baseline SARS-CoV-2 PCR-positive nasal swab), split approximately evenly across two age categories (ie, younger adults 18 to 64 years and older adults ≥65 years of age), yielding a subset not representative of the study’s age group distribution. The intentional “oversampling” of older adult participants was designed to provide a robust assessment of immunogenicity in this age group in whom SARS-CoV-2 infection is associated with higher risk of severe disease [19] and in whom immune responses may be blunted. Each age cohort was comprised of ≈400 participants who received NVX-CoV2373 and ≈200 who received placebo, reflecting the 2:1 randomization ratio.

In addition, we performed an ad hoc analysis of humoral immune responses in a subset of participants without evidence of previous SARS-CoV-2 infection who received NVX-CoV2373 (N = 119) and self-reported as people living with HIV(PLWH), and whose HIV infection was well-controlled by stable antiretroviral therapy (HIV RNA <50 copies/mL, CD4+ count >200 cells/mL). Their results were compared with those from a subset of participants without evidence of previous SARS-CoV-2 infection who were HIV-negative (N = 536).

### Study Procedures

Blood samples to measure serum antibody responses to the primary set of vaccinations on Days 0 and 21 were collected at baseline (before the first vaccination at Day 0) and at Day 35 (14 days following the second vaccination). Three assays were performed: immunoglobulin (IgG) antibodies to SARS-CoV-2 S protein, human angiotensin-converting enzyme 2 (hACE2) receptor binding inhibition antibodies to SARS-CoV-2 S protein, and neutralizing antibodies to SARS-CoV-2 wild-type virus (ancestral strain).

### Immunogenicity Assessments

Serum IgG antibody levels specific to SARS-CoV-2 S protein were measured using an enzyme-linked immunosorbent assay (ELISA) (Novavax Clinical Immunology, Gaithersburg, MD, USA), as described in Keech et al. [12]. The assay had a lower limit of quantification (LLOQ) of 200 ELISA units per mL.

hACE2 receptor binding inhibition antibodies to SARS-CoV-2 S protein were measured by ELISA (Novavax Clinical Immunology, Gaithersburg, MD, USA), as described by Tian et al [20]. The assay had an LLOQ of 10 inhibition antibody titers.

Neutralizing antibodies specific to SARS-CoV-2 were measured according to published methods [13] using a microneutralization assay with an inhibitory concentration of 50%, using wild-type virus strain SARS-CoV-2 hCoV-19/Australia/VIC01/2020 (GenBank MT007544.1) (360biolabs, Australia), which had an LLOQ of 20. For all 3 assays, titers below LLOQ were documented as 0.5 × LLOQ.

Evidence of previous exposure was determined at baseline by the presence of SARS-CoV-2 anti-nucleoprotein (NP) serum antibodies (University of Washington, Seattle, Washington, USA), using the Elecsys^®^ Anti-SARS-CoV-2 assay [21] and/or a positive SARS-CoV-2 Reverse Transcriptase Polymerase Chain Reaction (RT-PCR) in nasal swabs collected at baseline (University of Washington, Seattle, WA, USA) using the Abbott RealTime Quantitative SARS-CoV-2 Assay [17].

### Statistical Analyses

Two population analysis sets were analyzed. The per-protocol immunogenicity (PP-IMM) analysis set included participants previously unexposed to SARS-CoV-2 with a baseline and at least 1 serum sample result available after full primary vaccination and no major protocol violations considered relevant to the immune response. Participants were excluded from this analysis for nasal RT-PCR-positive swabs or anti-NP seropositivity for SARS-CoV-2 prior to the Day 35 visit. For samples collected on Day 35, participants must have received both vaccinations of the primary series within the specified 21 + ≈7-day window to be included in the PP-IMM analysis set. The second analysis set, per-protocol immunogenicity analysis set 2 (PP-IMM-2), was defined to evaluate the effect on immunogenicity of previous SARS-CoV-2 exposure. The PP-IMM-2 followed the same method described in the PP-IMM analysis set population, except that it included all participants regardless of baseline SARS-CoV-2 exposure status.

For assays detecting serum antibody levels specific to SARS-CoV-2 S protein antigen, the geometric mean, and geometric mean fold rise (GMFR) compared with baseline (Day 0), were summarized by trial vaccine group with 95% confidence interval (CI). The 95% CI was calculated based on the t-distribution of the log-transformed values for geometric means or GMFR, then back-transformed to the original scale for presentation. Seroconversion was a binary variable defined as 1 if the post-vaccination value was ≥4 times the baseline value.

Otherwise, the variable equaled 0. Seroconversion rate (SCR) was defined as a percentage of participants with a ≥4-fold higher antibody level on Day 35 vs Day 0. The 95% CI was calculated using the exact Clopper-Pearson method.

## RESULTS

### Study Population

Of 1200 participants randomly selected for the immunogenicity assessments, 1063 (88.6%) were in the PP-IMM analysis set, ie, participants without evidence of prior exposure to SARS-CoV-2 and without exclusionary protocol deviations, and 1119 (93.3%) were in the PP-IMM-2 analysis set, ie, participants with or without evidence of prior SARS-CoV-2 exposure and without exclusionary protocol deviations (Table 1 and Figure 1).

**Table 1.** Immunogenicity Analysis Sets (All Randomized Participants, Both Age Cohorts)

| Analysis Sets (Both Age Cohorts), n (%) | NVX-CoV2373<br>N = 805 | Placebo<br>N = 395 | Total<br>N = 1200 |
| --- | --- | --- | --- |
| Per-protocol immunogenicity analysis set | 717 (89.1) | 346 (87.6) | 1063 (88.6) |
| Per-protocol immunogenicity analysis set 2 | 759 (94.1) | 360 (86.2) | 1119 (93.3) |
| Baseline serologically/RT-PCR negative | 717 (94.5) | 346 (96.1) | 1063 (95.0) |
| Baseline serologically/RT-PCR positive | 42 (5.5) | 14 (3.9) | 56 (5.0) |
| 18–64 years | 26 (61.9) | 8 (57.1) | 34 (60.7) |
| ≥65 years | 16 (38.1) | 6 (42.9) | 22 (39.3) |

Abbreviations: RT-PCR, reverse transcriptase polymerase chain reaction.

**Figure 1.**
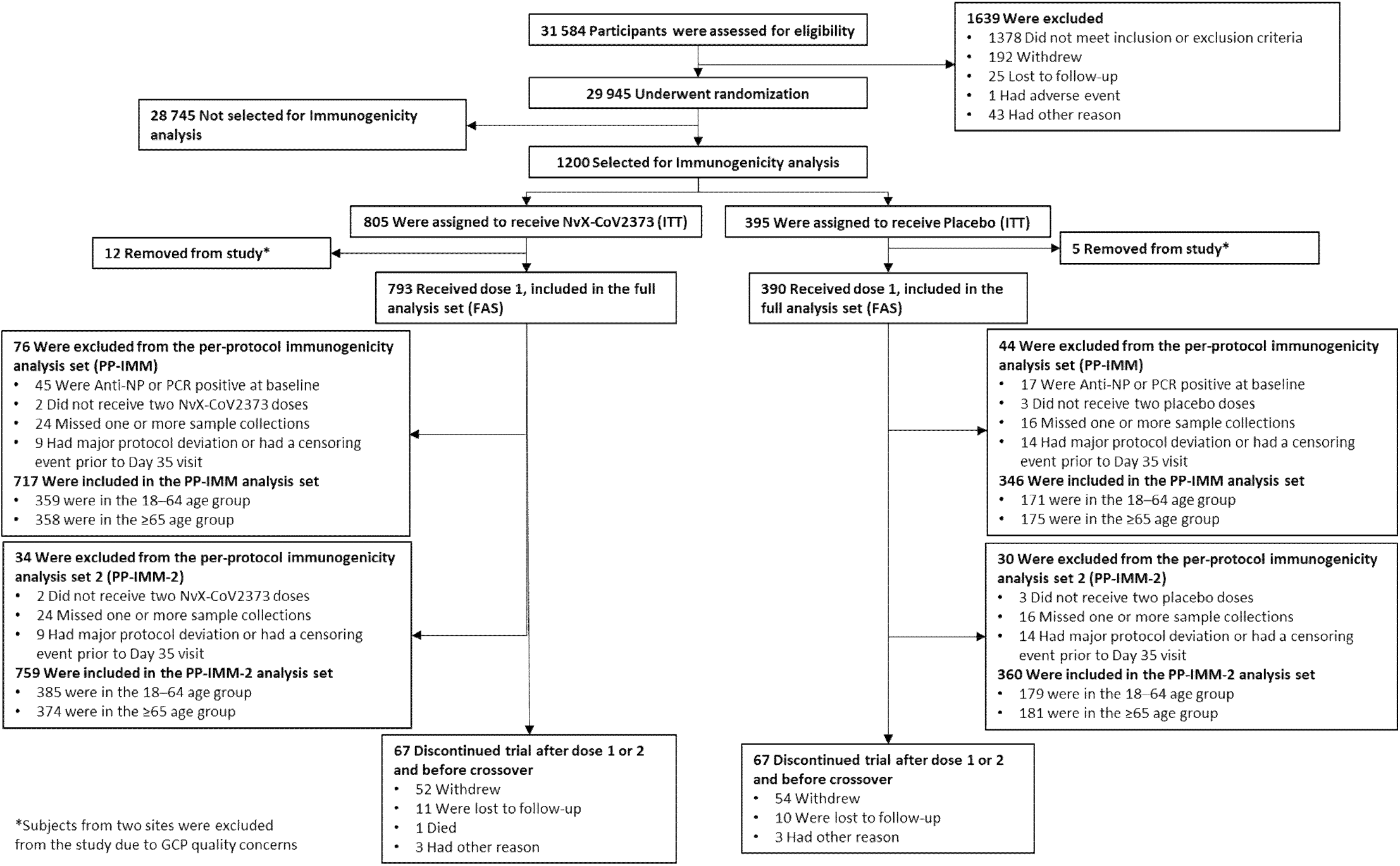
Distribution of participants by treatment groups and age cohorts. Abbreviations: FAS, full analysis set; GCP, good clinical practices; ITT, intent-to-treat; NP, nucleoprotein; PCR, Polymerase Chain Reaction; PP-IMM, Per-Protocol Immunogenicity analysis set; PP-IMM-2, Per-Protocol Immunogenicity analysis set 2.

Demographic characteristics of participants in the PP-IMM analysis set were well balanced between the two treatment groups (Table 2). Median age (range) was 65.0 years (18 to 95 years), with, as planned, approximately 50% of participants ≥65 years of age. Approximately half the participants were male (50.0%) and 19.2% were of Hispanic or Latino origin, while the majority were White (78.8%), and enrolled in the United States (93.1%). Participants from racial and ethnic minority groups were well represented and included those who were Black or African American (10.3%), American Indian or Native Alaskan (6.1%), and Asian (3.2%). In the Immunogenicity cohort, 56 participants (42 [5.5%] randomized to NVX-CoV2373; 26 in the 18 to 64 years group and 16 in the ≥65 years group, and 14 [3.9%] to placebo; 8 in the 18 to 64 years group and 6 in the ≥65 years group; Table 1 and Figure 1) were found retrospectively to be seropositive (ie, positive to SARS-CoV-2 anti-NP antibodies) or SARS-CoV-2 RT-PCR-positive at baseline. The overall baseline seropositivity/RT-PCR positivity in the PREVENT-19 trial was 6.6% [10]. Demographics of the PP-IMM and PP-IMM-2 by age cohorts can be found in Supplementary materials (Tables S1 to S4).

**Table 2.** Demographic Characteristics (Per-Protocol Immunogenicity Analysis Set, Both Age Cohorts)

| Parameter, n (%) | NVX-CoV2373<br>N = 717 | Placebo<br>N = 346 | Total<br>N = 1063 |
| --- | --- | --- | --- |
| Sex |  |  |  |
| Male | 366 (51.0) | 165 (47.7) | 531 (50.0) |
| Female | 351 (49.0) | 181 (52.3) | 532 (50.0) |
| Age (years) |  |  |  |
| Mean (SD) | 57.1 (15.73) | 56.6 (16.74) | 57.0 (16.06) |
| Median | 64.0 | 65.0 | 65.0 |
| Min–max | 18–95 | 18–87 | 18–95 |
| Age group |  |  |  |
| 18–64 years | 359 (50.1) | 171 (49.4) | 530 (49.9) |
| ≥65 years | 358 (49.9) | 175 (50.6) | 533 (50.1) |
| Race |  |  |  |
| White | 573 (79.9) | 265 (76.6) | 838 (78.8) |
| Black or African American | 62 (8.6) | 47 (13.6) | 109 (10.3) |
| American Indian or Alaska Native | 49 (6.8) | 16 (4.6) | 65 (6.1) |
| Native Hawaiian or Other Pacific<br>Islander | 0 | 1 (0.3) | 1 (<0.1) |
| Asian | 20 (2.8) | 14 (4.0) | 34 (3.2) |
| Mixed origin | 10 (1.4) | 2 (0.6) | 12 (1.1) |
| Not reported | 3 (0.4) | 1 (0.3) | 4 (0.4) |
| Ethnicity |  |  |  |
| Hispanic or Latino | 143 (19.9) | 61 (17.6) | 204 (19.2) |
| Not Hispanic or Latino | 573 (79.9) | 285 (82.4) | 858 (80.7) |
| Not reported | 1 (0.1) | 0 | 1 (<0.1) |
| Country |  |  |  |
| Mexico | 50 (7.0) | 23 (6.6) | 73 (6.9) |
| United States | 667 (93.0) | 323 (93.4) | 990 (93.1) |

Abbreviations: SD, standard deviation.

Mean (SD) baseline participant ages for HIV-positive and HIV-negative participants were 50.6 (12.2) years versus 44.8 (13.9) years, respectively. For HIV-positive versus HIV-negative participants, 81.51% versus 51.68% were male, 51.26% versus 83.02% were White, and 22.69% versus 16.60% were Hispanic/Latino, respectively.

### Immunogenicity Assessments

#### Neutralizing Antibodies

Neutralizing antibody geometric mean titers (GMTs) at Day 35 were increased in participants who were previously unexposed and received NVX-CoV2373 vs participants who received placebo in both age groups (Figure 2). At Day 0 (baseline), neutralizing antibody GMTs were near the LLOQ among participants in both treatment groups for each age group. At Day 35, neutralizing antibody GMTs in NVX-CoV2373 recipients were markedly increased relative to placebo recipients across the age groups (1302.7 vs 10.7 for younger adults and 899.8 vs 10.8 for older adults), with no evidence of a response in placebo recipients. Neutralizing antibody GMTs in the NVX-CoV2373 group were approximately 1.4-fold higher in the younger adults than in the older adults. These immune responses among vaccine recipients corresponded to neutralizing antibody GMFRs of 123.8 and 86.3 times higher than the baseline levels across the younger and older adults, respectively. Nearly all participants in both age groups seroconverted post-vaccination (98.3% for younger adults and 94.3% for the older adults).

**Figure 2.**
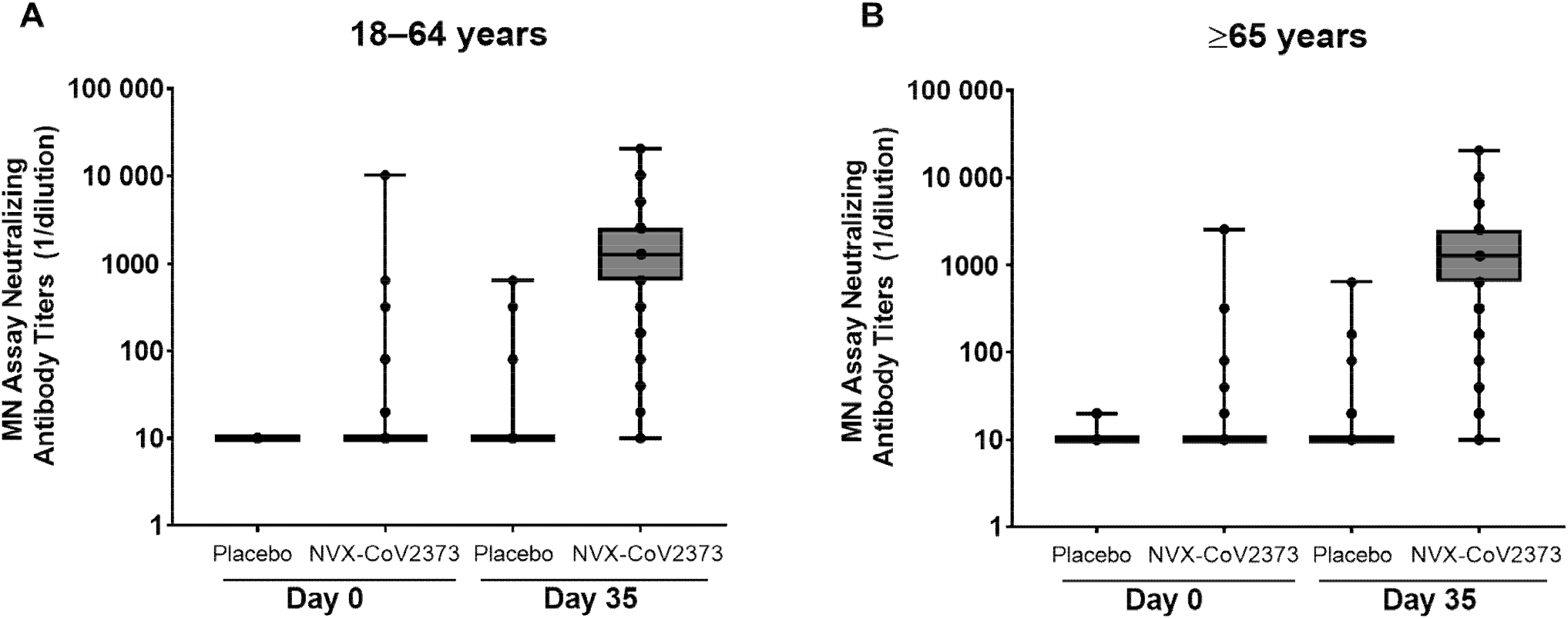
SARS-CoV-2 neutralizing antibody responses in participants who were seronegative/RT-PCR-negative at baseline. Panel A: participants 18–64 years, Panel B: participants ≥65 years. Abbreviations: MN, microneutralization; RT-PCR, reverse transcriptase polymerase chain reaction; SARS-CoV-2, severe acute respiratory syndrome coronavirus 2.

Participants who were seropositive at baseline randomized to either treatment were found to have quantifiable neutralizing antibodies at baseline. Among participants with evidence of prior exposure at baseline (both age groups combined), GMT at Day 35 were 3741.9 and 195.0 in the NVX-CoV2373 vs placebo groups, respectively (Table S5). GMT at Day 35 by age cohort are shown in Figure S1. Neutralizing antibody GMTs among NVX-CoV2373 recipients were higher at Day 35 in the previously exposed cohort compared with the previously unexposed cohort (Table S5).

#### Anti-S protein IgG antibodies

Serum IgG antibody levels to SARS-CoV-2 S protein 14 days after second vaccination (Day 35) among participants previously unexposed demonstrated a robust response compared with those who received placebo (Figure 3). At baseline (Day 0), serum IgG antibody geometric mean ELISA units (GMEUs) were near LLOQ for both treatment groups for each age group. At Day 35, serum IgG antibody GMEUs in the NVX-CoV2373 group were markedly higher than those observed in placebo recipients in both age groups (64 259.2 vs 120.9 for younger adults and 37 750.3 vs 132.7 for older adults) with no evidence of response among placebo recipients. Day 35 serum IgG antibody GMEUs in the NVX-CoV2373 group were approximately 1.7-fold higher in the younger adults (18 to 64 years of age) than in the older adults (≥65 years of age). Post-vaccination serum IgG antibody GMFRs were 549.8 and 310.7-fold over baseline in younger and older adults, respectively. Nearly all NVX-CoV2373 recipients seroconverted (98.1% for younger adults and 96.6% for older adults).

**Figure 3.**
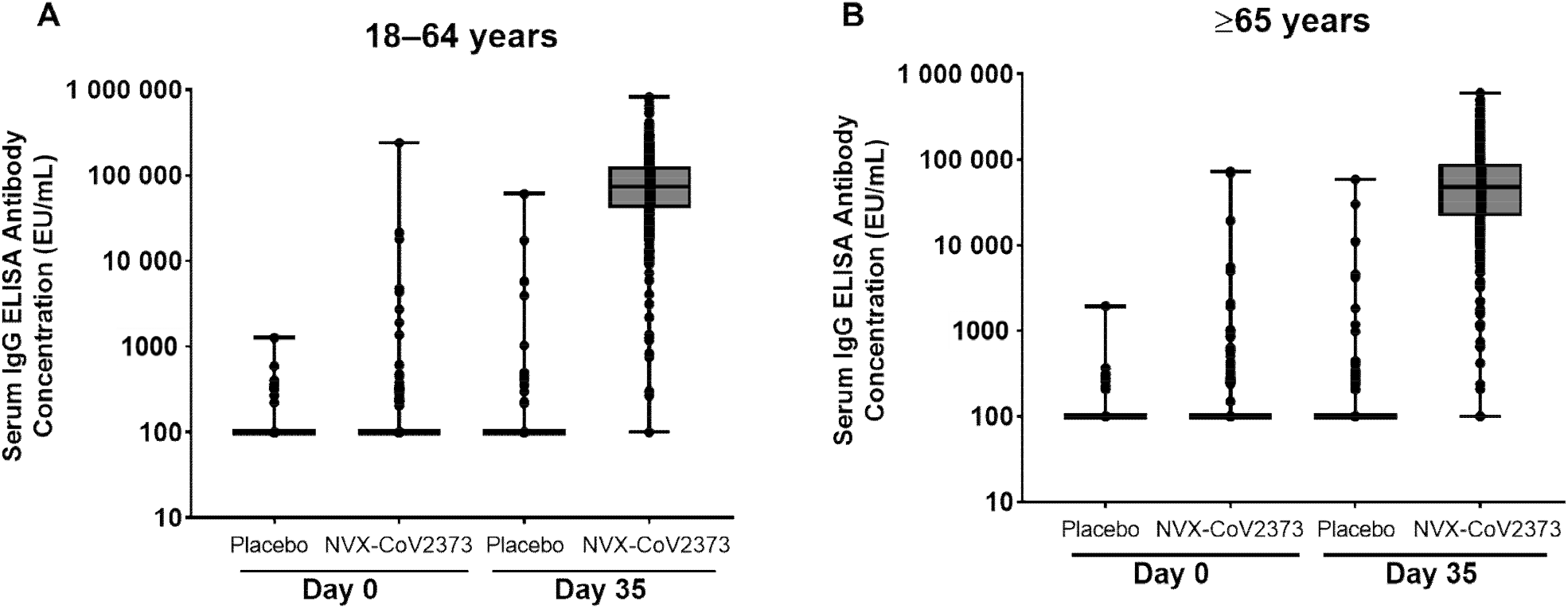
SARS-CoV-2 anti-Spike IgG antibody responses in participants who were baseline seronegative/RT-PCR-negative at baseline participants. Panel A: participants 18– to 64 years, Panel B: participants ≥ 65 years. Abbreviations: ELISA, enzyme-linked immunosorbent assay; EU/mL, ELISA units per mL; IgG, immunoglobulin; RT-PCR, reverse transcriptase polymerase chain reaction; SARS-CoV-2, severe acute respiratory syndrome coronavirus 2.

**Figure 4.**
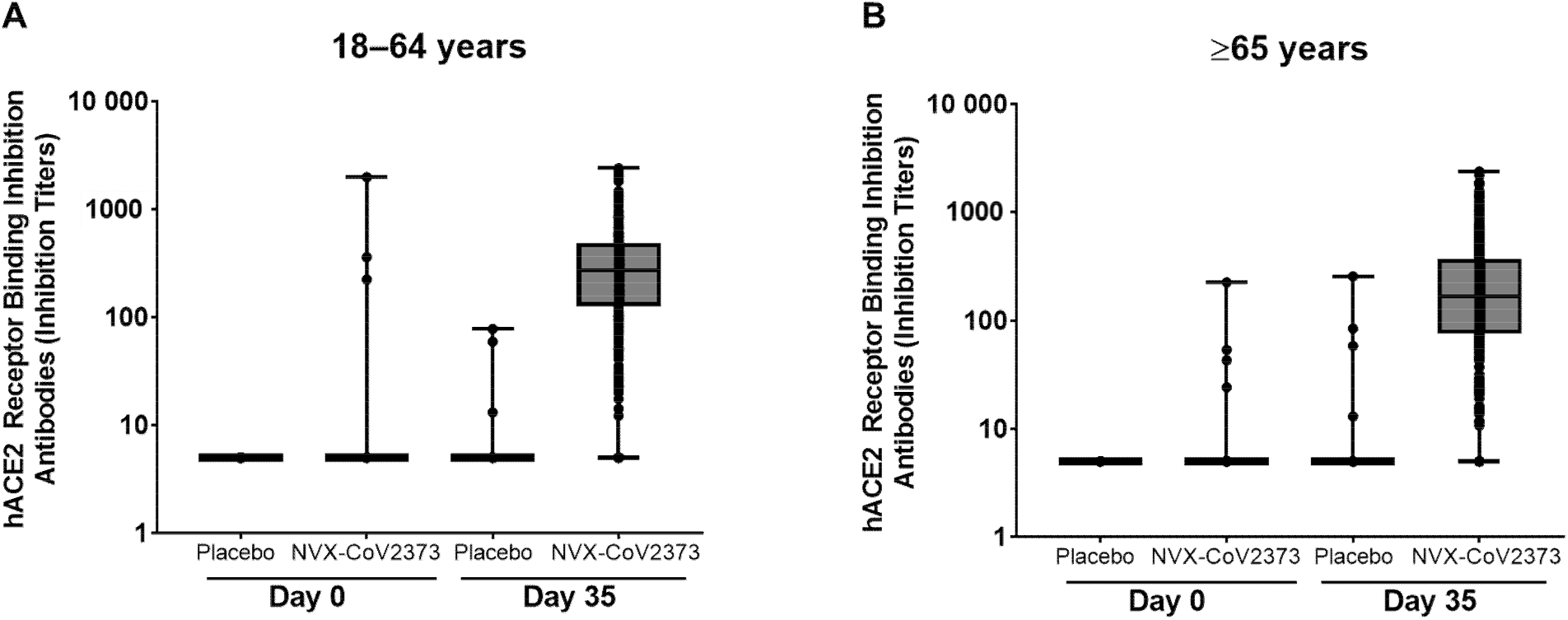
hACE2 receptor inhibition antibody responses in participants who were baseline seronegative/RT-PCR-negative at baseline participants. Panel A: participants 18 to 64 years, Panel B: participants ≥ 65 years. Abbreviations: hACE2, human angiotensin converting enzyme 2; RT-PCR, reverse transcriptase polymerase chain reaction.

Participants who were seropositive at baseline randomized to either treatment group were found to have, as expected, quantifiable anti-S IgG titers at baseline. Among participants with evidence of prior exposure at baseline (both age groups combined), GMEU at Day 35 were 119 620.4 and 6135.6 in the NVX-CoV2373 vs placebo groups, respectively (Table S6), and are shown by age cohort in Figure S2. A 2.4-fold higher serum IgG antibody response was observed when comparing GMEU from NVX-CoV2373 recipients who were previously exposed with those previously unexposed (119 620.4 vs 49 270.7, respectively; Table S6).

Serum IgG antibody levels were assessed in 119 PLWH who received NVX-CoV2373 (median age, 53 years; range 22–79 years) and who were anti-NP negative/RT-PCR negative at baseline; these levels were compared with those from a subset of 536 participants who were HIV-negative, healthy/medically stable, previously unexposed to SARS-CoV-2, and received NVX-CoV2373 (median age, 46 years; range, 18–78 years). Anti-S IgG antibodies demonstrated lower GMEU at Day 35 in PLWH who were slightly older (Table S7), compared with individuals who were HIV-negative (GMR: 0.63, 95% CI, 0.50–0.80) (Table 3); however, the GMFR in PLWH was 302-fold higher than baseline, confirming a robust response to the vaccine. Seroconversion rates were high (≥98.9%) and similar in PLWH and participants who were HIV-negative (difference 1.12%, 95% CI, −2.02 to 2.42).

**Table 3.**
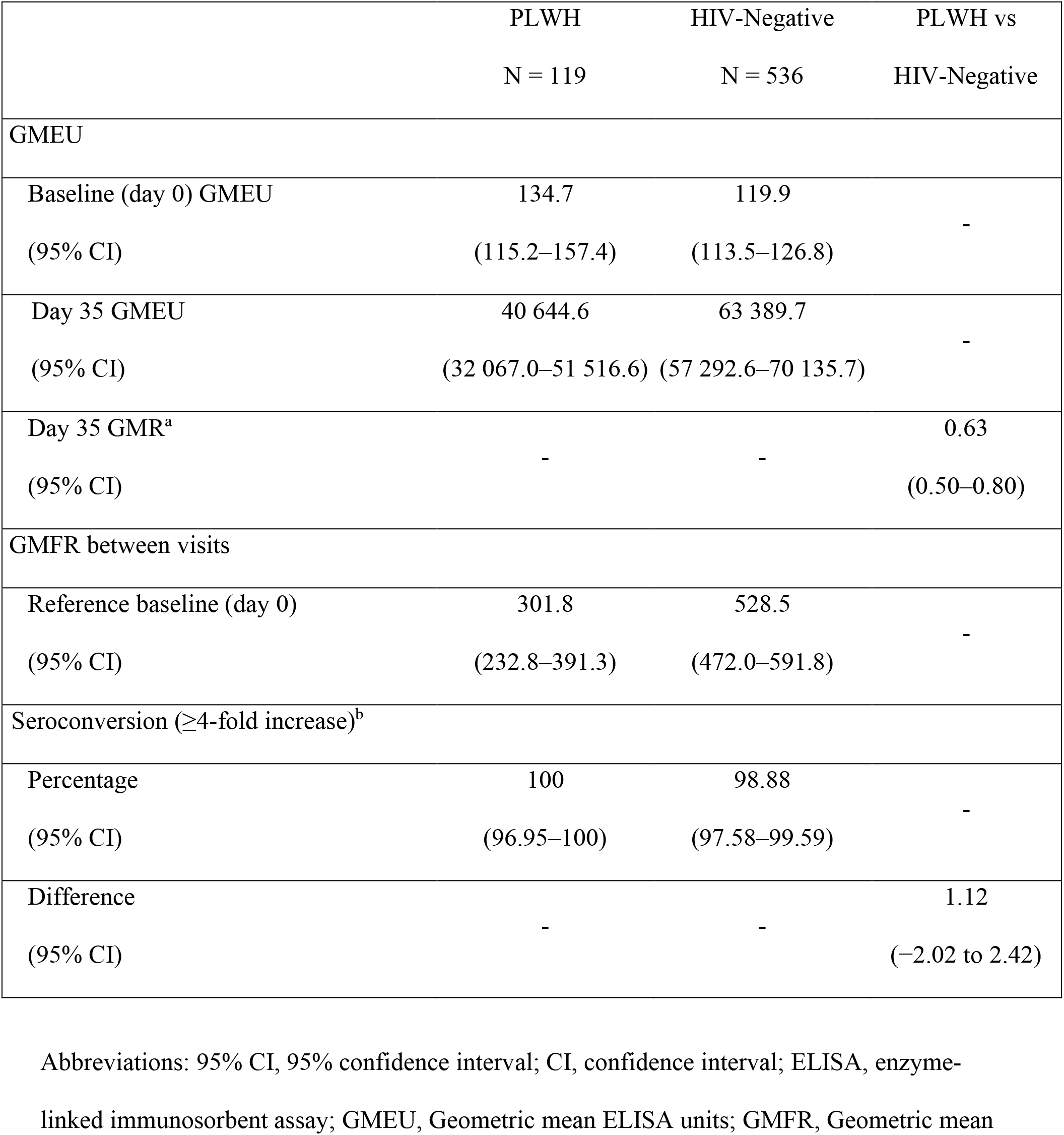
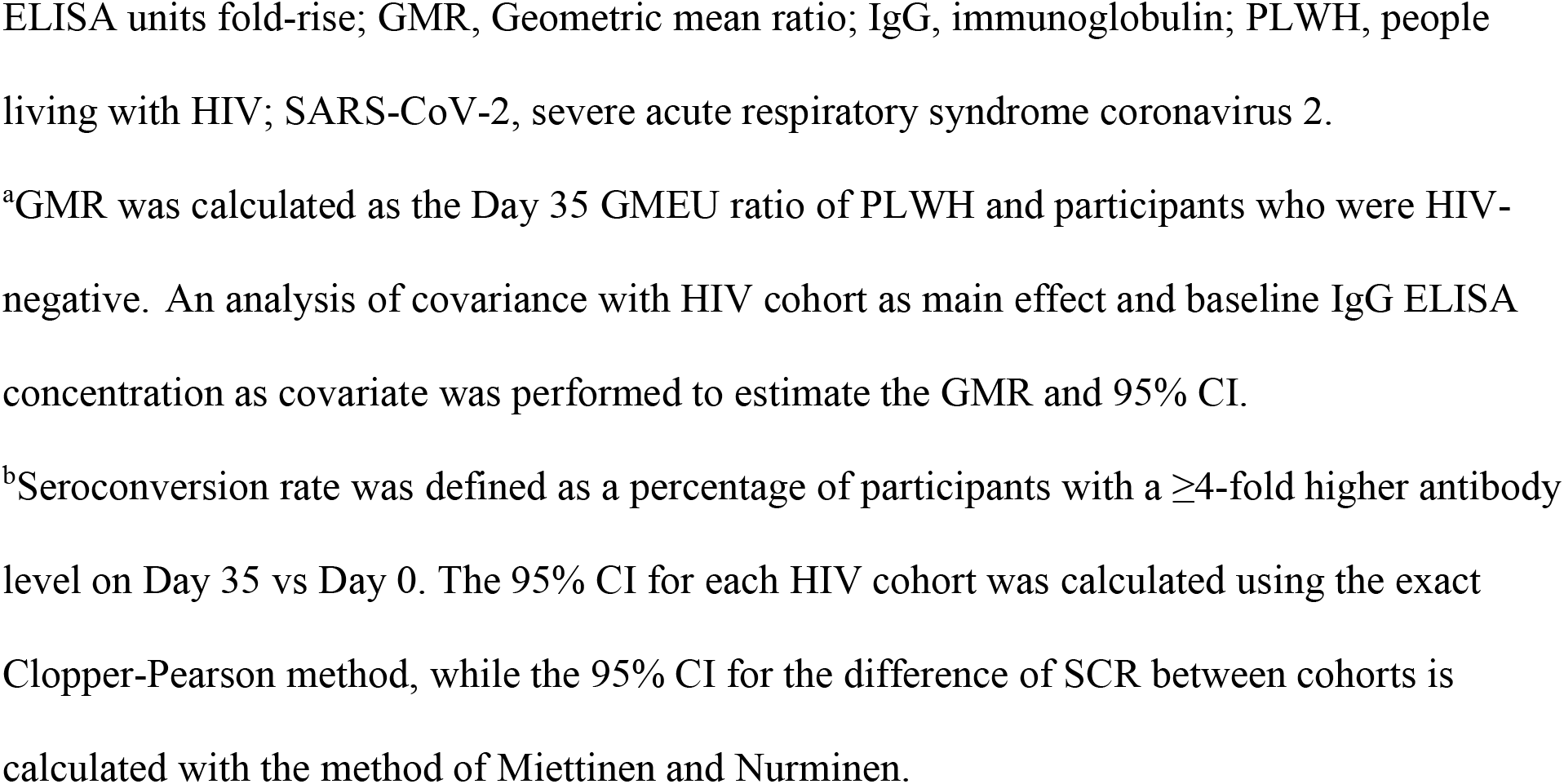
Serum Anti-Spike IgG Levels in PLWH and HIV-Negative Participants in PREVENT-19 Trial Who Received NVX-CoV2373, Without Evidence of Previous SARS-CoV-2 Infection.

|  | PLWH<br>N = 119 | HIV-Negative<br>N = 536 | PLWH vs<br>HIV-Negative |
| --- | --- | --- | --- |
| GMEU |  |  |  |
| Baseline (day 0) GMEU<br>(95% CI) | 134.7<br>(115.2–157.4) | 119.9<br>(113.5–126.8) | - |
| Day 35 GMEU<br>(95% CI) | 40 644.6<br>(32 067.0–51 516.6) | 63 389.7<br>(57 292.6–70 135.7) | - |
| Day 35 GMR<sup>a</sup><br>(95% CI) | - | - | 0.63<br>(0.50–0.80) |
| GMFR between visits |  |  |  |
| Reference baseline (day 0)<br>(95% CI) | 301.8<br>(232.8–391.3) | 528.5<br>(472.0–591.8) | - |
| Seroconversion ($\geq 4$-fold increase)<sup>b</sup> | | | |
| Percentage<br>(95% CI) | 100<br>(96.95–100) | 98.88<br>(97.58–99.59) | - |
| Difference<br>(95% CI) | - | - | 1.12<br>(–2.02 to 2.42) |

Abbreviations: 95% CI, 95% confidence interval; CI, confidence interval; ELISA, enzyme-linked immunosorbent assay; GMEU, Geometric mean ELISA units; GMFR, Geometric mean
ELISA units fold-rise; GMR, Geometric mean ratio; IgG, immunoglobulin; PLWH, people living with HIV; SARS-CoV-2, severe acute respiratory syndrome coronavirus 2.
<sup>a</sup>GMR was calculated as the Day 35 GMEU ratio of PLWH and participants who were HIV-negative. An analysis of covariance with HIV cohort as main effect and baseline IgG ELISA concentration as covariate was performed to estimate the GMR and 95% CI.
<sup>b</sup>Seroconversion rate was defined as a percentage of participants with a $\geq 4$ -fold higher antibody level on Day 35 vs Day 0. The 95% CI for each HIV cohort was calculated using the exact Clopper-Pearson method, while the 95% CI for the difference of SCR between cohorts is calculated with the method of Miettinen and Nurminen.

#### hACE2 Inhibition Antibodies

At baseline, hACE2 receptor binding inhibition antibody GMTs were near LLOQ across both treatment groups for each age group. At Day 35, hACE2 receptor binding inhibition antibody GMTs in NVX-CoV2373 recipients were markedly increased relative to in placebo recipients in both age groups (221.8 vs 5.2 for younger adults and 136.2 vs 5.3 for older adults), with no evidence of response among placebo recipients (Figure 3). hACE2 receptor binding inhibition antibody GMTs in the NVX-CoV2373 group were approximately 1.6-fold higher in the younger adults compared with the older adults. The hACE2 receptor binding inhibition GMFRs among NVX-CoV2373 recipients were 42.7 and 26.5 higher at Day 35 compared with baseline in the 2 age groups, respectively. SCRs in the NVX-CoV2373 recipients also were markedly increased across both age groups (95.3% for younger adults and 88.5% for older adults). Similar to what was observed for other assays, hACE2 receptor binding inhibition antibody GMTs at Day 35 were higher among participants with evidence of prior exposure at baseline (Table S8).

## DISCUSSION

The adjuvanted prefusion, stabilized, full length, recombinant SARS-CoV-2 S protein nanoparticle vaccine (NVX-CoV2373) containing the saponin-based adjuvant Matrix-M™ induced robust anti-S IgG antibodies, functional antibodies that block binding of SARS-CoV-2 spike protein to the hACE2 receptor, and neutralized the virus in adults aged ≥18 years. High antibody levels were observed in all three assays 14 days after the second dose of vaccine in both younger and older adults regardless of prior exposure status. These data confirm and expand upon previous immunogenicity results of NVX-CoV2373 in a smaller sample of adults in the phase 1 trial in US and Australia [13]. As expected, and as reported in the phase 1 and 2 studies [12, 13], humoral responses were higher among NVX-CoV2373 recipients who were previously exposed than those unexposed. Together with the earlier phase 2 and 3 trials [15, 16], the immunogenicity from PREVENT-19 with robust immune responses was consistent with the previously reported high VE for the prevention of COVID-19 due to a variety of circulating SARS-CoV-2 variants (VE = 90.4%, 95% confidence interval, CI: 82.9, 94.6) and for prevention of moderate-to-severe disease (VE = 100%, 95% CI: 87.0, 100) [17].

To date, serological thresholds predictive of VE against COVID-19 have not been defined fully. An analysis of correlates of immune responses with VE in the PREVENT-19 trial found that the anti-S IgG antibodies and neutralizing antibodies were each inversely correlated with risk of COVID-19 [22]. Additional studies to better define immune correlates of efficacy are ongoing. It is notable that neutralizing antibodies did not explain the observed VE of NVX-CoV2373 completely [22]. The potential contributions to NVX-CoV2373-induced protection by other functional responses, such as Fc-receptor functional antibodies [23] and T cell responses [20], are being explored.

The ability of COVID-19 vaccines to protect older adults is of great interest because of the risk of more severe disease complications in this population [19]. The sample size in the immunogenicity subset in our trial was enriched for older adults (≥65 years of age) to rigorously assess immunogenicity in this group. As expected, immune responses in older adults were approximately 40% lower for serum IgG and hACE2 receptor binding inhibition antibodies and approximately 30% for neutralizing antibodies, regardless of baseline serostatus. Nonetheless, SCRs were high and similar across the two age groups. Although the number of events was insufficient to assess VE in older adults participating in PREVENT-19, it is reassuring that the phase 3 trial of NVX-CoV2373 in the United Kingdom demonstrated 88.9% VE (95% CI: 20.2, 99.7) among adults 65–84 years of age [16]. The main variant circulating in the United Kingdom at the time of the study was Alpha, as was the case in PREVENT-19 [17].

PLWH are considered at increased risk of developing severe illness from COVID-19 [19]. Less robust IgG responses have been observed in PLWH in other COVID-19 vaccine trials, including a trial of NVX-CoV2373 conducted in South Africa [15, 24]. In most studies, the lower response rates appeared to be associated with lower CD4+ T cell counts, but most PLWH ultimately seroconverted [25]. The GMEUs among PLWH in this trial, in whom older age may have contributed to lower immune responses, were similar to those associated with a high degree of protective efficacy in an earlier study of NVX-CoV2373 [26].

Several limitations of this study are worth noting. This subset was intentionally selected to be split nearly evenly across the 2 age categories. Thus, the selected immunogenicity subset is not representative of the total enrolled study population in which over 85% of the population were in the 18–64 age group. Moreover, the median age of the PP-IMM analysis set was 65 years compared with 47 years for the PP-EFF analysis set. Therefore, a combined analysis of immunogenicity that includes both age cohorts cannot be used to correlate immunogenicity with the overall VE results reported previously [17]. In addition, the number of NVX-CoV2373 recipients who were previously exposed to SARS-CoV-2 available for assessment of humoral responses in naturally infected individuals (ie, anti-NP seropositive/RT-PCR positive at baseline) was limited, given that previous presence of laboratory confirmed COVID-19 was exclusionary from the trial. Whether these immune responses are generalizable to current and future variants will require continued monitoring of effectiveness over time.

In conclusion, NVX-CoV2373 elicited robust immune responses measured by anti-S specific IgG antibody binding levels, as well as by functional assays (ie, neutralizing antibody and hACE2 receptor binding inhibition antibody) at 2 weeks following second dose in the primary vaccination series. Seroconversion rates were high and comparable across both age groups; however, titers were higher in younger adults compared with older adults. Humoral responses in participants who were baseline seropositive/RT-PCR positive were, as expected, higher than in participants who were seronegative/RT-PCR negative. NVX-CoV2373 induced a robust anti-S IgG response in HIV-positive individuals comparable to that induced by other SARS-CoV-2 vaccines in PLWH [27] and may be expected to provide satisfactory protection against COVID-19 in this population.

Additional durability of immune responses after primary vaccination and effects of booster vaccinations for ancestral/variant SARS-CoV-2 will be reported soon (unpublished data).

## Supporting information

CONSORT Checklist

## Data Availability

Study information is available at https://clinicaltrials.gov/ct2/show/NCT04611802 and requests will be considered.

## Acknowledgments

We especially thank all the study participants who volunteered for the study, as well as to the 2019nCoV-301 Study Group members (listed in the Supplementary materials); to the funders (Biomedical Advanced Research and Development Authority, National Institute of Allergy and Infectious Diseases (NIAID)/National Institutes of Health), the members of the NIAID Data and Safety Monitoring Board, community leadership groups throughout the country who assisted with community engagement and recruitment; and unnamed colleagues at each of the sites who generously contributed to the trials in many ways, and all unnamed colleagues at Novavax, Inc., who worked tirelessly and gave unlimited efforts to the development, testing, and support of this trial. Editorial assistance on the preparation of this manuscript was provided by Kelly M. Cameron, of Ashfield MedComms, an Inizio company, supported by Novavax, Inc.

## Author contributions

Conceived and designed the activity: GÁ, LMD

Acquired data: KLK, CLG, JN, HD

Analyzed: GÁ, LMD, SC-C, AMG, WW, IC, JSP

Interpreted the data: GÁ, KLK, CLG, AMG, WW, IC, GMG, LMD

Wrote first draft of the manuscript: GÁ

Approved final version of the manuscript: All authors

