## Appendix for "Immunogenicity of NVX-CoV2373 in PREVENT-19: A Phase 3, Randomized, Placebo-Controlled Trial in Adults in the United States and Mexico"

### Supplementary Material

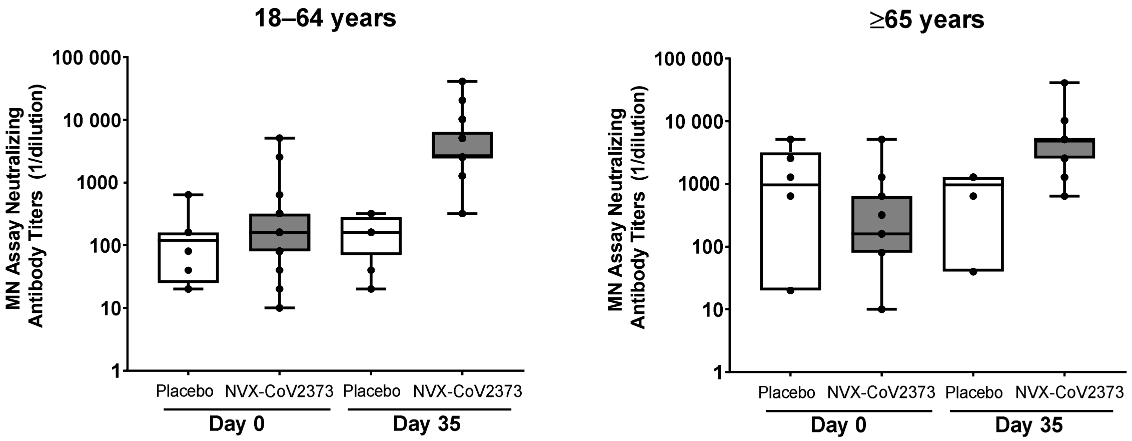


**Figure S1.** SARS-CoV-2 neutralizing antibody responses in participants who were seropositive/RT-PCR-positive at baseline (PP-IMM-2 Analysis Set). Panel A: participants 18 to 64 years, Panel B: participants ≥ 65 years. Abbreviations: MN, microneutralization; PP-IMM, Per-Protocol Immunogenicity analysis set; RT-PCR, reverse transcriptase polymerase chain reaction; SARS-CoV-2, severe acute respiratory syndrome coronavirus 2.

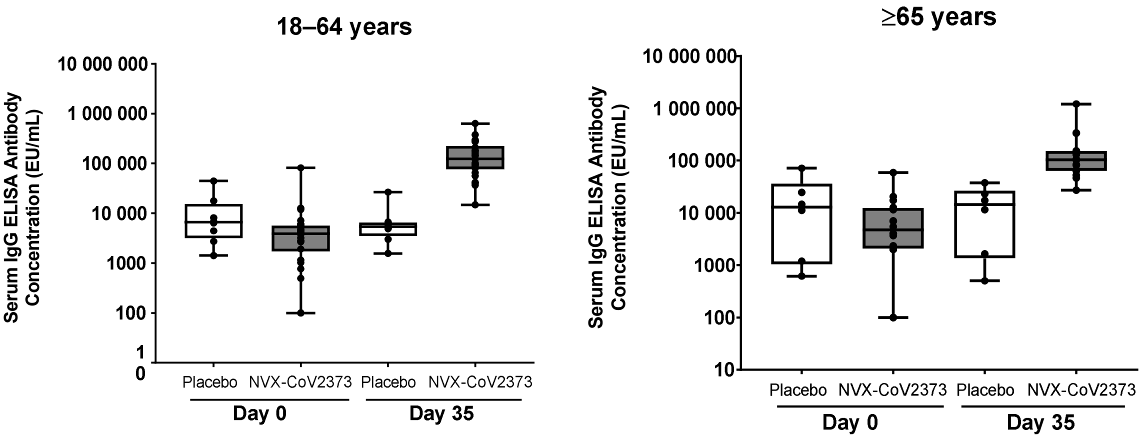


**Figure S2.** SARS-CoV-2 anti-spike IgG antibody responses in participants who were seropositive/RT-PCR-positive at baseline (PP-IMM-2 Analysis Set). Panel A: participants 18 to 64 years, Panel B: participants ≥ 65 years. Abbreviations: ELISA, enzyme-linked immunosorbent assay; EU/mL, ELISA units per mL; IgG, immunoglobulin; RT-PCR, reverse transcriptase polymerase chain reaction; SARS-CoV-2, severe acute respiratory syndrome coronavirus 2.

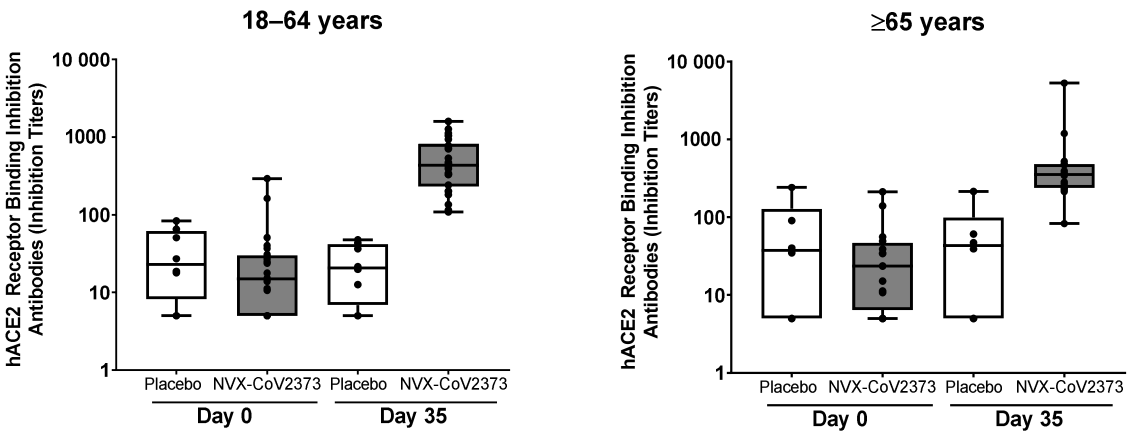

**Figure S3.** hACE2 receptor inhibition antibody responses in participants who were seropositive/RT-PCR-positive at baseline (PP-IMM-2 Analysis Set). Panel A: participants 18 to 64 years, Panel B: participants ≥ 65 years. Abbreviations: hACE2, human angiotensin converting enzyme 2; RT-PCR, reverse transcriptase polymerase chain reaction.

**Table S1. Demographic Characteristics (Per-Protocol Immunogenicity Analysis Set, 18 to 64 Years)**

| Parameter, n (%) | NVX-CoV2373<br>N = 359 | Placebo<br>N = 171 | Total<br>N = 530 |
|---|---|---|---|
| Sex | | | |
| Male | 185 (51.5) | 80 (46.8) | 265 (50.0) |
| Female | 174 (48.5) | 91 (53.2) | 265 (50.0) |
| Age (years) | | | |
| Mean (SD) | 44.6 (12.33) | 42.7 (12.79) | 43.9 (12.50) |
| Median | 46.0 | 43.0 | 45.0 |
| Min–max | 18–64 | 18–64 | 18–64 |
| Race | | | |
| White | 289 (80.5) | 129 (75.4) | 418 (78.9) |
| Black or African American | 31 (8.6) | 22 (12.9) | 53 (10.0) |
| American Indian or Alaska Native | 17 (4.7) | 5 (2.9) | 22 (4.2) |
| Native Hawaiian or Other Pacific Islander | — | 1 (0.6) | 1 (0.2) |
| Asian | 13 (3.6) | 13 (7.6) | 26 (4.9) |
| Mixed Origin | 7 (1.9) | 1 (0.6) | 8 (1.5) |
| Not Reported | 2 (0.6) | — | 2 (0.4) |
| Ethnicity | | | |
| Hispanic or Latino | 82 (22.8) | 34 (19.9) | 116 (21.9) |

| Parameter, n (%) | NVX-CoV2373<br><br>N = 359 | Placebo<br><br>N = 171 | Total<br><br>N = 530 |
|---|---|---|---|
| Not Hispanic or Latino | 276 (76.9) | 137 (80.1) | 413 (77.9) |
| Not Reported | 1 (0.3) | — | 1 (0.2) |
| Country | | | |
| Mexico | 18 (5.0) | 8 (4.7) | 26 (4.9) |
| United States | 341 (95.0) | 163 (95.3) | 504 (95.1) |

Abbreviations: SD, standard deviation.

**Table S2. Demographic Characteristics (Per-Protocol Immunogenicity Analysis Set, ≥65 Years)**

| Parameter, n (%) | NVX-CoV2373<br>N = 358 | Placebo<br>N = 175 | Total<br>N = 533 |
|---|---|---|---|
| Sex | | | |
| Male | 181 (50.6) | 85 (48.6) | 266 (49.9) |
| Female | 177 (49.4) | 90 (51.4) | 267 (50.1) |
| Age (years) | | | |
| Mean (SD) | 69.8 (4.93) | 70.3 (4.20) | 69.9 (4.70) |
| Median | 68.0 | 69.0 | 69.0 |
| Min–max | 65–95 | 65–87 | 65–95 |
| Race | | | |
| White | 284 (79.3) | 136 (77.7) | 420 (78.8) |
| Black or African American | 31 (8.7) | 25 (14.3) | 56 (10.5) |
| American Indian or Alaska Native | 32 (8.9) | 11 (6.3) | 43 (8.1) |
| Native Hawaiian or Other Pacific Islander | — | — | — |
| Asian | 7 (2.0) | 1 (0.6) | 8 (1.5) |
| Mixed Origin | 3 (0.8) | 1 (0.6) | 4 (0.8) |
| Not Reported | 1 (0.3) | 1 (0.6) | 2 (0.4) |
| Ethnicity | | | |
| Hispanic or Latino | 61 (17.0) | 27 (15.4) | 88 (16.5) |

| Parameter, n (%) | NVX-CoV2373<br><br>N = 358 | Placebo<br><br>N = 175 | Total<br><br>N = 533 |
|---|---|---|---|
| Not Hispanic or Latino | 297 (83.0) | 148 (84.6) | 445 (83.5) |
| Not Reported | 1 (0.3) | 1 (0.6) | 2 (0.4) |
| Country | | | |
| Mexico | 32 (8.9) | 15 (8.6) | 47 (8.8) |
| United States | 326 (91.1) | 160 (91.4) | 486 (91.2) |

Abbreviations: SD, standard deviation.

**Table S3. Demographic Characteristics (Per-Protocol Immunogenicity Analysis Set 2, 18 to 64 Years)**

| Parameter, n (%) | NVX-CoV2373<br>N = 385 | Placebo<br>N = 179 | Total<br>N = 564 |
|---|---|---|---|
| Sex | | | |
| Male | 197 (51.2) | 83 (46.4) | 280 (49.6) |
| Female | 188 (48.8) | 96 (53.6) | 284 (50.4) |
| Age (years) | | | |
| Mean (SD) | 44.6 (12.39) | 42.5 (12.83) | 43.9 (12.56) |
| Median | 46.0 | 43.0 | 45.0 |
| Min–max | 18–64 | 18–64 | 18–64 |
| Race | | | |
| White | 303 (78.7) | 132 (73.7) | 435 (77.1) |
| Black or African American | 41 (10.6) | 24 (13.4) | 65 (11.5) |
| American Indian or Alaska Native | 19 (4.9) | 7 (3.9) | 26 (4.6) |
| Native Hawaiian or Other Pacific Islander | — | 1 (0.6) | 1 (0.2) |
| Asian | 13 (3.4) | 14 (7.8) | 27 (4.8) |
| Mixed Origin | 7 (1.8) | 1 (0.6) | 8 (1.4) |
| Not Reported | 2 (0.5) | — | 2 (0.4) |
| Ethnicity | | | |
| Hispanic or Latino | 96 (24.9) | 36 (20.1) | 132 (23.4) |

| Parameter, n (%) | NVX-CoV2373<br><br>N = 385 | Placebo<br><br>N = 179 | Total<br><br>N = 564 |
|---|---|---|---|
| Not Hispanic or Latino | 288 (74.8) | 143 (79.9) | 431 (76.4) |
| Not Reported | 1 (0.3) | — | 1 (0.2) |
| Country | | | |
| Mexico | 22 (5.7) | 10 (5.6) | 32 (5.7) |
| United States | 363 (94.3) | 169 (94.4) | 532 (94.3) |

Abbreviations: SD, standard deviation.

**Table S4. Demographic Characteristics (Per-Protocol Immunogenicity Analysis Set 2, ≥65 Years)**

| Parameter, n (%) | NVX-CoV2373<br>N = 374 | Placebo<br>N = 181 | Total<br>N = 555 |
|---|---|---|---|
| Sex | | | |
| Male | 189 (50.5) | 88 (48.6) | 277 (49.9) |
| Female | 185 (49.5) | 93 (51.4) | 278 (50.1) |
| Age (years) | | | |
| Mean (SD) | 69.7 (4.85) | 70.2 (4.18) | 69.9 (4.65) |
| Median | 68.0 | 69.0 | 69.0 |
| Min–max | 65–95 | 65–87 | 65–95 |
| Race | | | |
| White | 294 (78.6) | 139 (76.8) | 433 (78.0) |
| Black or African American | 35 (9.4) | 26 (14.4) | 61 (11.0) |
| American Indian or Alaska Native | 34 (9.1) | 13 (7.2) | 47 (8.5) |
| Native Hawaiian or Other Pacific Islander | — | — | — |
| Asian | 7 (1.9) | 1 (0.6) | 8 (1.4) |
| Mixed Origin | 3 (0.8) | 1 (0.6) | 4 (0.7) |
| Not Reported | 1 (0.3) | 1 (0.6) | 2 (0.4) |
| Ethnicity | | | |
| Hispanic or Latino | 64 (17.1) | 30 (16.6) | 94 (16.9) |

| Parameter, n (%) | NVX-CoV2373<br><br>N = 374 | Placebo<br><br>N = 181 | Total<br><br>N = 555 |
|---|---|---|---|
| Not Hispanic or Latino | 310 (82.9) | 151 (83.4) | 461 (83.1) |
| Not Reported | — | — | — |
| Country | | | |
| Mexico | 35 (9.4) | 17 (9.4) | 52 (9.4) |
| United States | 339 (90.6) | 164 (90.6) | 503 (90.6) |

Abbreviations: SD, standard deviation.

**Table S5. Neutralizing Antibodies for SARS-CoV-2 Wild-Type Virus at Day 0 (Baseline) and Day 35 (14 Days After Second Vaccination) in Adult Participants by Baseline Serostatus (PP-IMM-2 Analysis Set)**

| Parameter | Serologically/RT-PCR Negative | | Serologically/RT-PCR Positive | |
|---|---|---|---|---|
| | NVX-CoV2373 N = 717 | Placebo N = 346 | NVX-CoV2373 N = 42 | Placebo N = 14 |
| Day 0 (baseline) | | | | |
| n1 | 714 | 344 | 41 | 14 |
| GMT | 10.5 | 10.1 | 160.0 | 168.1 |
| 95% CI[a] | 10.2, 10.9 | 10.0, 10.1 | 97.4, 262.9 | 55.7, 507.1 |
| Day 35 | | | | |
| n1 | 709 | 345 | 42 | 14 |
| GMT | 1082.9 | 10.8 | 3741.9 | 195.0 |
| 95% CI[a] | 973.0, 1205.3 | 10.2, 11.3 | 2754.9, 5082.5 | 87.0, 437.1 |

Abbreviations: CI, confidence interval; GMT, geometric mean titer; n1, number of participants in the PP-IMM-2 Analysis Set with non-missing data at visit; PP-IMM-2, Per Protocol Immunogenicity 2; RT-PCR, reverse transcriptase polymerase chain reaction.

[a]The 95% CI for GMT were calculated based on the t-distribution of the log-transformed values, then back transformed to the original scale for presentation.

**Table S6. Serum IgG Antibody Concentrations to SARS-CoV-2 S Protein at Day 0 (Baseline) and Day 35 (14 Days After Second Vaccination) in Adult Participants by Baseline Serostatus (PP-IMM-2 Analysis Set)**

| Parameter | Serologically/RT-PCR Negative | | Serologically/RT-PCR Positive | |
|---|---|---|---|---|
| | NVX-CoV2373 N = 717 | Placebo N = 346 | NVX-CoV2373 N = 42 | Placebo N = 14 |
| Day 0 (baseline) | | | | |
| n1 | 717 | 345 | 42 | 14 |
| GMEU | 119.2 | 108.2 | 3062.2 | 7212.4 |
| 95% CI[a] | 112.8, 125.9 | 104.5, 112.1 | 1859.6, 5042.6 | 3253.5, 15 988.5 |
| Day 35 | | | | |
| n1 | 717 | 344 | 42 | 14 |
| GMEU | 49 270.7 | 126.7 | 119 620.4 | 6135.6 |
| 95% CI* | 44 521.3, 54 526.8 | 115.6, 139.0 | 91 715.7, 156 015.1 | 3042.4, 12 373.9 |

Abbreviations: CI, confidence interval; ELISA, enzyme-linked immunosorbent assay; GMEU, geometric mean ELISA units; IgG, immunoglobulin G; n1, number of participants in the PP-IMM-2 Analysis Set with non-missing data at visit; PP-IMM-2, Per Protocol Immunogenicity 2; RT-PCR, reverse transcriptase polymerase chain reaction.

[a]The 95% CI for GMEU were calculated based on the t-distribution of the log-transformed values, then back transformed to the original scale for presentation.

**Table S7. Demographic Characteristics of Participants in PREVENT-19 Who Were HIV-Positive and HIV-Negative, and Seronegative/RT-PCR Negative to SARS-CoV-2 at Baseline, Who Received 2 Doses of NVX-CoV2373**

| Characteristic, n (%) | NVX-CoV2373 Recipients | | |
|---|---|---|---|
| | HIV-Positive<br>n = 119 | HIV-Negative<br>n = 536 | Total<br>N = 655 |
| Age (years) | | | |
| Median (range) | 53 (22–79) | 46 (18–78) | 48 (18–79) |
| Age group | | | |
| 18 to 64 years | 107 (89.9) | 509 (95.0) | 616 (94.0) |
| ≥ 65 years | 12 (10.1) | 27 (5.0) | 39 (6.0) |
| Sex | | | |
| Male | 97 (81.5) | 277 (51.7) | 374 (57.0) |
| Female | 22 (18.5) | 259 (48.3) | 281 (43.0) |
| Race | | | |
| White | 61 (51.3) | 445 (83.0) | 506 (77.3) |
| Black or African American | 44 (37.0) | 52 (9.7) | 96 (14.7) |
| American Indian or Alaska Native | 2 (1.7) | 2 (0.4) | 4 (0.6) |
| Asian | 3 (2.5) | 25 (4.7) | 28 (4.3) |
| Multiple | 7 (5.9) | 9 (1.7) | 16 (2.4) |

| Characteristic, n (%) | NVX-CoV2373 Recipients | | |
|---|---|---|---|
| | HIV-Positive<br>n = 119 | HIV-Negative<br>n = 536 | Total<br>N = 655 |
| Native Hawaiian or Other Pacific Islander | 1 (0.8) | 2 (0.4) | 3 (0.5) |
| Not reported | 1 (0.8) | 1 (0.2) | 2 (0.3) |
| Ethnicity | | | |
| Not Hispanic or Latino | 91 (76.5) | 446 (83.2) | 537 (82.0) |
| Hispanic or Latino | 27 (22.7) | 89 (16.6) | 116 (17.7) |
| Not reported or Unknown | 1 (0.8) | 1 (0.2) | 2 (0.3) |

Abbreviations: RT-PCR, reverse transcriptase polymerase chain reaction; SARS-CoV-2, severe acute respiratory syndrome coronavirus 2.

**Table S8. hACE2 Receptor Binding Inhibition Antibodies to SARS-CoV-2 S Protein at Day 0 (Baseline) and Day 35 (14 Days After Second Vaccination) in Adult Participants by Baseline Serostatus (PP-IMM-2 Analysis Set)**

| Parameter | Serologically/RT-PCR Negative | | Serologically/RT-PCR Positive | |
|---|---|---|---|---|
| | NVX-CoV2373<br>N = 717 | Placebo<br>N = 346 | NVX-CoV2373<br>N = 42 | Placebo<br>N = 14 |
| Day 0 (baseline) | | | | |
| n1 | 717 | 346 | 42 | 14 |
| GMT | 5.2 | 5.0 | 18.2 | 25.4 |
| 95% CI[a] | 5.0, 5.3 | 5.0, 5.0 | 12.8, 25.8 | 12.3, 52.6 |
| Day 35 | | | | |
| n1 | 716 | 345 | 42 | 14 |
| GMT | 173.9 | 5.2 | 411.2 | 21.9 |
| 95% CI[a] | 157.8, 191.7 | 5.0, 5.4 | 320.0, 528.3 | 11.2, 42.9 |

Abbreviations: CI, confidence interval; GMT, geometric mean titer; hACE2, human angiotensin-converting enzyme 2; n1, number of participants in the PP-IMM-2 Analysis Set with non-missing data at visit; PP-IMM-2, Per Protocol Immunogenicity 2; RT-PCR, reverse transcriptase polymerase chain reaction; SARS-CoV-2, severe acute respiratory syndrome coronavirus 2

[a]The 95% CI for GMT were calculated based on the t-distribution of the log-transformed values, then back transformed to the original scale for presentation.

**Table S9. 2019nCoV-301 Study Group (in Alphabetical Order of Institution Affiliation)**

| Affiliation/Funding* | Study Group | Location |
|---|---|---|
| México | | |
| Centro de Atención e Investigación Médica (CAIMED) | Jorge F. Méndez Galván, MD, Monica B. Carrascal, Adriana Sordo Duran, Laura Ruy Sanchez Guerrero, Martha Cecilia Gómora Madrid | Mexico City, Mexico |
| FAICIC Clinical Research | Alejandro Quintín Barrat Hernández, MD, Sharzhaad Molina Guizar, Denisse Alejandra González Estrada, Silvano Omar Martínez Pérez, MD, Zindy Yazmín Zárate Hinojosa, MD | Veracruz, Mexico |
| Instituto Nacional de Ciencias Médicas y Nutrición Salvador Zubirán | Guillermo Miguel Ruiz-Palacios, MD | Mexico City, Mexico |
| Instituto Nacional de Salud Pública | Aurelio Cruz-Valdez, PhD, Janeth Pacheco-Flores, MD, Anyela Lara, MD, Secia Díaz-Miralrio | Cuernavaca, Mexico |
| PanAmerican Clinical Research México | María José Reyes Fentanes, MD, Jocelyn Zuleica Olmos Vega, MD, Daniela Pineda Méndez, MD, Karina Cano Martínez, MD, Winniberg Stephany Alvarez León | Querétaro, Mexico |
| PanAmerican Clinical Research México | Vida Veronica Ruiz Herrera, MD, Eduardo Gabriel Vázquez Saldaña, Laura Julia Camacho Choza, Karen Sofia Vega Orozco, Sandra Janeth Ortega Domínguez | Guadalajara, Mexico |
| Unidad de Atención Médica e Investigación en Salud (UNAMIS) | Jorge A. Chacón, MD, Juan J. Rivera, MD, Erika A. Cutz, MD, Maricruz E. Ortegón, MD, María I. Rivera, MD | Mérida, Mexico |
| United States and Puerto Rico | | |
| Accellacare | David Browder, MD, Cortney Burch, Terri Moye, Paul Bondy, MD, Lesley Browder, MD | Rocky Mount, NC |
| Accellacare | Rickey D. Manning, MD, James Wilson Hurst, MD, Rodney E. Sturgeon, MD, Paul H. Wakefield, MD, John A. Kirby, MD | Knoxville, TN |
| Accel Research Sites | James Andersen, MD, Szheckera Fearon, MSN, FNP-C, Rosa Negron, MD, Amy Medina, ADN, BS | Lakeland, FL |
| Accel Research Sites | Bruce Rankin, DO, John M. Hill, MD, Steven Shinn, MD, Vivek Rajasekhar, DO, Marshall Nash, MD | DeLand, FL |
| Achieve Clinical Research | Hayes Williams, MD, PhD, LaShondra Cade, Rhodna Fouts, Connie Moya | Birmingham, AL |
| Alliance for Multispecialty Research | Corey G. Anderson, MD, Naomi Devine, NP-C, James Ramsey, NP-C, Ashley Perez, David Tatelbaum | Tempe, AZ |
| Alliance for Multispecialty Research | Michael Jacobs, MD, Kathleen Menasche, LPN, Vincent Mirkil, MD | Las Vegas, NV |
| Anaheim Clinical Trials | Peter J. Winkle, MD, Amina Z. Haggag, MD, Michelle Haynes, Marysol Villegas, Sabina Raja | Anaheim, CA |
| Atlanta Center for Medical Research | Robert Riesenberg, MD, Stanford Plavin, MD, Mark Lerman, MD, Leana Woodside, DNP, NP-C, Maria Johnson, MD | Atlanta, GA |
| Baylor College of Medicine / NIAID (UM1AI148575) | C. Mary Healy, MD, Jennifer A. Whitaker, MD, Hana El Sahly, MD, Christine Akamine, MD, Wendy A. Keitel, MD, Robert L. Atmar, MD | Houston, TX |
| Biomedical Advanced Research and | Richard Gorman, MD, Gary Horwith, MD, Robin Mason, MS, MBA | Washington, DC |

| | | |
|---|---|---|
| Development Authority (BARDA) | | |
| Benchmark Research | Laurence Chu, MD, Michelle Chouteau, MD, Lisa Johnson, FNP, Tambra Dora | Austin, TX |
| Benchmark Research | Greg Hachigian, MD, Deborah Murray, FNP, Michael Cancilla, PA, Logan Ledbetter, PA, Masaru Oshita, MD | Sacramento, CA |
| Benchmark Research | William Seger, MD, Beverly Ewing, APRN, DNP, FNP-BC | Fort Worth, TX |
| Beth Israel Deaconess Medical Center / NIAID (UM1AI068614) | Kathryn E. Stephenson, MD, MPH, Chen Sabrina Tan, MD, Rebecca Zash, MD, Jessica L. Ansel, MSN, Kate Jaegle, MSN, Caitlin J. Guiney, MSN | Boston, MA |
| Black Hills Center for American Indian Health / Missouri Breaks Industries Research Inc / NIAID (UM1AI068614) | Jeffrey A. Henderson, MD, MPH, Marcia O'Leary, RN, Kendra Enright, RN, Jill Kessler, MS, Pete Ducheneaux, LPN, Asha Inniss, MS, APRN | Eagle Butte, SD |
| California Research Foundation | Donald M. Brandon, MD, William B. Davis, MD, Daniel T. Lawler, MD | San Diego, CA |
| Carolina Institute for Clinical Research | Yaa D. Oppong, MD, Ryan P. Starr, DO, Scott N. Syndergaard, DO, Rozeli Shelly, MD, Mashrur Islam Majumder | Fayetteville, NC |
| Cedar Crosse Research Center | Danny Sugimoto, MD, Jeffrey Dugas Sr., MD, Dolores Rijos, Sandra Shelton, Stephan Hong, MD | Chicago, IL |
| Cenexel RCA | Howard Schwartz, MD, Nelia Sanchez-Crespo, MD, Jennifer Schwartz, APRN, Terry Piedra, BS, Barbara Corral, APRN | Hollywood, FL |
| Centex Studies | Joel Solis, MD, Carmen Medina, PA, Westley Keating, PA | McAllen, TX |
| Clinical Neuroscience Solutions | Michael E. Dever, MD, Mitul Shah, MD, Michael Delgado, MD, Tameika Scott, DrPH | Orlando, FL |
| Clinical Neuroscience Solutions | Lisa S. Usdan, MD, Lora J. McGill, MD, Valerie K. Arnold, MD, Carolyn Scatamacchia, MSN, NP-C, Codi M. Anthony, DNP, APRN, PMHNP-BC | Memphis, TN |
| CommonSpirit Health Research Institute | Rajan Merchant, MD, Anelgine Crans Yoon, MD, Janet Hill, PA-C, Lucy Ng-Price, MA, Teri Thompson-Seim | Woodland, CA |
| Comprehensive Clinical Research | Ronald Ackerman, MD, Jamie Ackerman, Florida Aristy, APRN | West Palm Beach, FL |
| Covid-19 Prevention Network (CoVPN) | Lawrence Corey, MD, Kathleen M Neuzil, MD, MPH, Huub G Gelderblom, MD, PhD, Nzeera Ketter, Carrie Sopher | Seattle, WA |
| CRA Headlands | Jon Finley, MD, Nathan Segall, MD, Mildred Stull, APRN, FNP-C | Stockbridge, GA |
| DM Clinical Research | Vicki E. Miller, MD, MPH, Monica Murray, Blanca Gomez, Zainab Rizvi, Sonia Guerrero | Tomball, TX |
| Empire Clinical Research | Yogesh K. Paliwal, MD, Amit Paliwal, MD, Sarah Gordon, MS, Bryan Gordon, Cynthia Montano-Pereira | Pomona, CA |
| Headlands Research | Christopher Galloway, MD, Candice Montros, Lily Aleman, Samira Shairi, RN, Wesley Van Ever | Orlando, FL |
| Health Research of Hampton Roads | George H. Freeman, MD, Esther Laverne Harmon, ANP, Marshall A. Cross, MD, Kacie Sales, BSN, RN, Catherine Q. Gular, PharmD | Newport News, VA |

| | | |
|---|---|---|
| HHS-DoD Countermeasures Acceleration Group | Matthew Hepburn, MD | Washington, DC |
| HOPE Research Institute | Matthew Doust, MD, Nathan Alderson, PhD, Shana Harshell | Phoenix, AZ |
| Howard University Hospital / Howard University College of Medicine / NIAID (UM1AI068614) | Siham Mahgoub, MD, Celia Maxwell, MD, Thomas Mellman, MD, Karl M Thompson, PhD, Glenn Wortman, MD | Washington, DC |
| IACT Health | Jeff Kingsley, DO, April Pixler, LaKondria Curry, Sarah Afework, Austin Swanson | Columbus, GA |
| Jacksonville Center for Clinical Research | Jeffry Jacqmein, MD, Maggie Bowers, PA-C, Dawn Robison, APRN-C, Victoria Mosteller, MD, Janet Garvey, DNP | Jacksonville, FL |
| Johnson County Clin-Trials | Carlos Fierro, MD, Mary Easley, BSN, RN | Lenexa, KS |
| Joint Program Executive Office for Chemical, Biological, Radiological and Nuclear Defense's, US Department of Defense | Rebecca J. Kurnat | Washington, DC |
| Lynn Health Science Institute | Carl P. Griffin, MD, Raymond Cornelison, MD, Shanda Gower, APRN, CNP, William Schnitz, MD, Destiny S. Heinzig-Cartwright, BA | Oklahoma City, OK |
| Lynn Institute of the Ozarks | Derek Lewis, MD, Fred E. Newton, MD, Aeiress Duhart, Breana Watkins, Brandy Ball | Little Rock, AR |
| Lynn Institute of the Rockies | Ripley Hollister, MD, Jeremy Brown, DO, Melody Ronk, PA-C, Jill York, Shelby Pickle | Colorado Springs, CO |
| M3-Emerging Medical Research | David B. Musante, MD, William P. Silver, MD, Linda R. Belhorn, MD, Nicholas A. Viens, MD, David Dellaero, MD | Durham, NC |
| M3-Wake Research | Matthew Hong, MD, Wayne Harper, MD, Lisa Cohen, DO, Priti Patel, NP, Kendra Lisec, PA | Raleigh, NC |
| MD Clinical | Beth Safirstein, MD, Luz Zapata, MD, Lazaro Gonzalez, APRN, Evelyn Quevedo, APRN, Farah Irani, PhD | Hallandale Beach, FL |
| Medical Research International | Joseph Grillo, MD, Amy Potts, PA-C, MPH, Julie White, MBA | Oklahoma City, OK |
| Medical University of South Carolina | Patrick Flume, MD, Gary Headden, MD, Brandie Taylor, NP, Ashley Warden, Amy Chamberlain | Charleston, SC |
| MedPharmics | Robert Jeanfreau MD, Susan Jeanfreau MD | Metairie, LA |
| MedPharmics | Paul G. Matherne, MD, Amy Caldwell, RN, Jessica Stahl, Mandy Vowell, Lauren Newhouse | Gulfport, MS |
| Meharry Medical College / NIAID (UM1AI068614) | Vladimir Berthaud MD, MPH, Zudi-Mwak Takizala MD, MPH, MBA, Genevieve Beninati, FNP, Kimberly Snell, PharmD, Sherrie Baker, BS, James Walker, RN | Nashville, TN |
| Meridian Clinical Research | David Ensz, MD, Tavane Harrison, CNP, Meagan Miller, Janet Otto | Sioux City, IA |
| Meridian Clinical Research | Brandon Essink, MD, Roni Gray, APRN, Christine Wilson, Tiffany Nemecek, Hannah Harrington, MPH | Omaha, NE |
| Meridian Clinical Research | Charles Harper, MD, Keith Vrbicky, MD, Chelsie Nutsch, NP, Sally Eppenbach, NP, Wendell Lewis, NP | Norfolk, NE |

| | | |
|---|---|---|
| Meridian Clinical Research | Jordan Whatley, MD, Christopher Dedon, APRN, FNP-C, Tana Bourgeois, RN, Lyndsea Folsom, Crystal Rowell, APRN, FNP-C | Baton Rouge, LA |
| Miami Veterans Affairs Medical Center / NIAID (UM1AI068614) | Gregory Holt, MD, Mehdi Mirsaeidi, MD, Rafael Calderon, MD, Paola Lichtenberger, MD, Jalima Quintero, RN, Becky Martinez, RN | Miami, FL |
| Morehouse School of Medicine / NIAID (UM1AI068614) | Lilly Immergluck, MD, Erica Johnson, PhD, Austin Chan, MD, Norberto Fas, MD, LaTeshia Thomas-Seaton, MS, APRN, Saadia Khizer, MD, MPH | Atlanta, GA |
| MultiCare Institute for Research and Innovation | Jonathan Staben, MD | Cheney, WA |
| National Institute of Allergy and Infectious Diseases (NIAID) / National Institutes of Health (NIH) | Tatiana Beresnev, MD, Maryam Jahromi, MD, Mary A. Marovich, MD, Julia Hutter, MD, Martha Nason, PhD, Julie Ledgerwood, DO, John Mascola, MD | Bethesda, MD |
| National Research Institute | Mark Leibowitz, MD, Fernanda Morales, Mike Delgado, Rosario Sanchez, Norma Vega | Los Angeles, CA |
| Novavax, Inc. | Lisa M. Dunkle, MD, Germán Áñez, MD, Gary Albert, Erin Coston, Chinar Desai, Haoua Dunbar, Mark Eickhoff, Jenina Garcia, Margaret Kautz, Angela Lee, Maggie Lewis, Alice McGarry, Irene McKnight, Joy Nelson, Patrick Newingham, Patty Price-Abbott, Patty Reed, Diana Vegas, Bethanie Wilkinson, PhD, Katherine Smith, MD, Wayne Woo, MS, Iksung Cho, MS, Gregory M. Glenn, MD, Filip Dubovsky, MD, MPH | Gaithersburg, MD |
| Omega Medical Research | David L. Fried, MD, Lynne A. Haughey, MSN, FNP, Ariana C. Stanton, PA-C, Lisa Stevens Rameaka, MD | Warwick, RI |
| Pharmacology Research Institute | David Rosenberg, MD, Lee Tomatsu, Viviana Gonzalez, Millie Manalo | Los Alamitos, CA |
| PMG Research of Bristol | Bernard Grunstra, MD, Donald Quinn, MD, Phillip Claybrook, MD, Shelby Olds, MD, Amy Dye | Bristol, TN |
| PMG Research of Wilmington | Kevin D. Cannon, MD, Mesha M. Chadwick, MD, Bailey Jordan, Morgan Hussey, Hannah Nevarez | Wilmington, NC |
| Ponce de Leon Center / NIAID (UM1AI068614) | Colleen F. Kelley, MD, MPH, Valeria D. Cantos MD, Michael Chung MD, Caitlin Moran, MD, MSc, Paulina Rebolledo, MD, Christina Bacher, PAC | Atlanta, GA |
| Ponce School of Medicine / NIAID (UM1AI148685) | Elizabeth Barranco-Santana, MD, Jessica Rodriguez, MD, Rafael Mendoza, MD, Karen Ruperto, MD, Odette Olivieri, MD, Enrique Ocaña, MD | Ponce, Puerto Rico |
| Preferred Research Partners | Paul E. Wylie, MD, Renea Henderson, DO, Natasa Jenson, MD, Fan Yang, MD, Amy Kelley, BSN, RN | Little Rock, AR |
| Providea Health Partners Elligo Health Research | Kenneth Finkelstein, DO, David Beckmann, MD, Tanya Hutchins, FNP, Sebastian Garcia Escallon, BA, Kristen Johnson | Evergreen Park, IL |
| Providence Clinical Research | Teresa S. Sligh, MD, Parul Desai, NP, Vincent Huynh, BSc, Carlos Lopez, MD, Erika Mendoza, BA | North Hollywood, CA |
| Research Your Health | Jeffrey Adelglass, MD, Jerome (Jerry) G. Naifeh, MD, Kristine Jane Kucera, PA-C, MPAS, DHS, Waseem Chughtai, BS, MBBS, Shireen Hasham Jaffer | Plano, TX |

| | | |
|---|---|---|
| Rochester Clinical Research | Matthew G. Davis, MD, Jennifer Foley, Michelle Lyn Burgett, RN, Tammi Louise Shlotzhauer, MD, Sarah Michelle Ingalsbe-Geno, RPA-C | Rochester, NY |
| SIMEDHealth / SIMEDResearch | Daniel Duncanson, MD, Kelly Kush, Lori Nesbitt, Cora Sonnier, Jennifer McCarter | Gainesville, FL |
| Sterling Research Group | Michael B. Butcher, MD, James Fry, PA-C, Donna Percy, RN, BSN, Karen Freudemann | Cincinnati, OH |
| Sterling Research Group | Bruce C. Gebhardt, MD, Padma N. Mangu, MD, Debra Beck Schroeck, MS, PA-C, Rajesh Kumar Davit, MD, Gayle D. Hennekes, PA-C, MPAS | Cincinnati, OH |
| Stony Brook University - Stony Brook Medicine / NIAID (UM1AI068614) | Benjamin J. Luft, MD, Melissa Carr, BA, Sharon Nachman, MD, Alison Pellecchia, BA, Candace Smith, PharmD, Bruno Valenti, NP | Commack, NY |
| Suncoast Research Associates | Maria I. Bermudez, MD, Noris Peraita, ARNP, Ernesto Delgado, ARNP, Alicia Arrazcaeta, Natalie Ramirez | Miami, FL |
| Suncoast Research Group | Mark E. Kutner, MD, Jorge Caso, MD, Janet Mendez, ARNP, Marianela Carvajal, ARNP, Carmen Amador, ARNP | Miami, FL |
| Sundance Clinical Research | Larkin Tyler Wadsworth III, MD, Horacio Marafioti, MD, Lyly Dang, DNP-BC, Lauren Clement, NP-C, Jennifer Berry, FNP-BC | St. Louis, MO |
| Synexus Clinical Research | Mohammed Allaw, MD, Georgettea Geuss, Chelsea Miles, NP, Zachary Bittner, Melody Werne | Evansville, IN |
| Synexus Clinical Research | Cornell Calinescu, MD, Shannon Rodman, Joshua Rindt | Henderson, NV |
| Synexus Clinical Research | Erin Cooksey, MD, Kristina Harrison, Deanna Cooper, Manisha Horton<br>Amanda Philyaw | Anderson, SC |
| Synexus Clinical Research | William Jennings, MD, Hilario Alvarado, MD, Michele Baka, MD, Malina Regalado, NP | San Antonio, TX |
| Synexus Clinical Research | Linda Murray, DO | Pinellas Park, FL |
| Synexus Clinical Research | Sherif Naguib, MD, Justin Singletary, Sha-Wanda Richmond, Sarah Omodele, Emily Oppenheim | Atlanta, GA |
| Synexus Clinical Research | Joseph Newberg, MD, Laura Pearlman, MD, Reuben Martinez, Victoria Andriulis | Chicago, IL |
| Synexus Clinical Research | Paul J. Nugent, DO, Leonard Singer, MD, Jeanne Blevins, Meagan Thomas, Christine Hull | Cincinnati, OH |
| Synexus Clinical Research | Isabel Pereira, MD, Gina Rivero, Tracy Okonya, Frances Downing, Paulina Miller | Vista, CA |
| Synexus Clinical Research | Margaret Rhee, MD, Katherine Stapleton, Jeffrey Klein, Rosamond Hong, MD | Akron, OH |
| Synexus Clinical Research | Suzanne Swan, MD, Tami Wahlin, MD, Elizabeth Bennett, PA, Amy Salzl<br>Sharine Phan | Richfield, MN |
| Synexus Clinical Research | Jewel Johnny White, MD, Amanda Occhino, Ruth Paiano APRN, Morgan McLaughlin APRN, Elisa Swieboda APRN | The Villages, FL |
| Texas Center for Drug Development | Veronica Garcia-Fragoso, MD, Maria Gabriela Becerra, MD, Cecilia Mckeown, Lisa Holloway, Toni White | Houston, TX |
| The Charlotte-Mecklenburg Hospital Authority d/b/a Atrium | Christine B. Turley, MD, Andrew McWilliams, MD, Tiffany Esinhart, PA-C, Natasha Montoya, APRN, Shamika Huskey, FNP, Leena Paul, FNP | Charlotte, NC |

| | | |
|---|---|---|
| Health / NIAID (UM1AI068614) | | |
| The Miriam Hospital / NIAID (UM1AI068636) | Karen Tashima, MD, Jennie Johnson, MD, Marguerite Neill, MD, Martha Sanchez, MD, Natasha Rybak, MD, Maria Mileno, MD | Providence, RI |
| UC Davis Health / NIAID (UM1AI068614) | Stuart H. Cohen, MD, Monica Ruiz, Dean M. Boswell, BS, Elizabeth E. Robison, BS, Trina L. Reynolds, BS, Sonja Neumeister, MPH | Sacramento, CA |
| Universidad de Puerto Rico - Recinto de Ciencias Médicas - Maternal Infant Studies Center (CEMI) / NIAID (UM1AI068636) | Carmen D. Zorrilla, MD, Juana Rivera, MD, MPH, Jessica Ibarra, MD, Iris García, BSN, RN, Dianca Sierra, BA, Wanda Ramon, BSPh | San Juan, Puerto Rico |
| University of Colorado Hospital CRS / NIAID (UM1AI068636) / NCATS (UL1TR002535, UM1AI069432) | Thomas B. Campbell, MD, Suzanne Fiorillo, MSPH, Rebecca Pitotti, RNP, Victoria Riedel Anderson, MS, Jose Castillo Mancilla, MD, Nga Le, PharmD | Aurora, CO |
| University of Iowa Medical Center / NIAID (UM1AI068614) / NCATS (UL1TR002537) | Patricia L. Winokur, MD, Dilek Ince, MD, Theresa Hegmann, PA, Jeffrey Meier, MD, Jack Stapleton, MD, Laura Stulken, PA | Iowa City, IA |
| University of Maryland School of Medicine / NIAID (UM1AI148689) | Monica McArthur, MD, PhD, Karen L. Kotloff, MD, Kathleen Neuzil, MD, Andrea Berry, MD, Milagritos Tapia, MD, Elizabeth Hammershaimb, MD, MS, Toni Robinson, RN, Rosa MacBryde, RN | Baltimore, MD |
| University of Minnesota / NIAID (UM1AI068614) | Susan Kline, MD, MPH, Joanne L. Billings, MD, MPH, Winston Cavert, MD,<br>Les B. Forgosh. MD, Timothy W. Schacker, MD, Tyler D. Bold, MD, PhD | Minneapolis, MN |
| University of Missouri Health Care / NIAID (UM1AI148685) | Dima Dandachi, MD, MPH, Taylor Nelson, DO, Andres Bran, MD,<br>Grant Geiger, S. Hasan Naqvi, MD | Columbia, MO |
| University of Nebraska Medical Center / NIAID (UM1AI068614) | Diana F Florescu, MD, Richard Starlin, MD, David Kline, MD, Andrea Zimmer, MD, Anum Abbas, MD, Natasha Wilson, APRN | Omaha, NE |
| University of North Carolina / NIAID (UM1AI068619) / University of North Carolina at Chapel Hill Center for AIDS Research (P30AI050410) / NC TraCS Institute (UL1TR002489) | Cynthia L. Gay, MD, MPH, Joseph J Eron, MD, Michael Sciaudone, MD, MPH, A. Lina Rosengren, MD, MPH, MS, Arianne Morrison, MD, Sarah E Rutstein, MD, PhD, Michael Herce, MD, MPH, Michelle Flores-Moore, MD, MS | Chapel Hill, NC |
| University of South Florida, Morsani College of Medine / NIAID (UM1AI068614) | Carina A. Rodriguez, MD, Elizabeth Bruce, MD, Claudia Espinosa, MD, Lisa J Sanders, MD, Kami Kim, MD, Denise Casey, RN | Tampa, FL |
| University of Texas Health Science Center San Antonio / NIAID (UM1AI068614) | Barbara S. Taylor, MD, MS, Thomas Patterson, MD, Ruth Serrano Pinilla, MD, Delia Bullock, MD, Philip Ponce, MD, Jan Patterson, MD | San Antonio, TX |

| | | |
|---|---|---|
| University of Washington / Lummi Tribal Health Center / NIAID (UM1AI148573) | R. Scott McClelland, MD, MPH, Dakotah C. Lane, MD, Anna Wald, MD, MPH, Frank James, MD, Elizabeth Duke, MD, Kirsten Hauge, MPH, Jessica Heimonen, MPH | Seattle, WA |
| University of Washington | Robert W. Coombs, MD, PhD, Alex Greninger, MD, PhD, MS, MPhil, Pavitra Roychoudhury, PhD, Erin A. Goecker, MS, Yunda Huang, PhD, Youyi Fong, PhD | Seattle, WA |
| VA Ann Arbor Healthcare System / NIAID (UM1AI068614) | Carol Kauffman, MD, Kathleen Linder, MD, Kimberly Nofz, BSN, Andrew McConnell, BS | Ann Arbor, MI |
| Velocity Clinical Research | Robert J. Buynak, MD, Angella Webb, APRN, Taryn Petty, FNP, Stephanie Andree, FNP | Valparaiso, IN |
| Velocity Clinical Research | Judith Kirstein, MD, Marcia Bernard, Erica Sanchez, Nolan Mackey, Clarisse Baudelaire | Banning, CA |
| Velocity Clinical Research | Gregg Lucksinger, MD, Jaleh Ostovar, NP | Medford, OR |
| Velocity Clinical Research | Mary Beth Manning, MD, Joan Rothenberg, MD, Toby Briskin, MD, Denise Roadman, PAC, Sarah Dzigiel | Cleveland, OH |
| Velocity Clinical Research | J. Scott Overcash, MD, Adrienna Marquez, Hanh Chu, Kia Lee, Kim Quillin | La Mesa, CA |
| Velocity Clinical Research | Barbara Rizzardi, MD, Michelle King, NP, Vanessa Abad, NP, Jennifer Knowles, BS | West Jordan, UT |
| Velocity Clinical Research | Michael Waters, MD, Karla Zepeda, NP, Scott Overcash, MD, Jordan Coslet, NP, Dalia Tovar, MA | Chula Vista, CA |
| Velocity Clinical Research | Marian E. Shaw, MD, Mark A. Turner, MD, Cory J. Huffine, FNP-C, Esther S. Huffine, FNP-C | Meridian, ID |
| Walter Reed Army Institute of Research | Julie A. Ake, MD, MSc | Silver Spring, MD |
| Wayne State University / NIAID (UM1AI068614) | Elizabeth Secord, MD, Eric McGrath, MD, Phillip Levy, MD, Brittany Stewart, RD, PharmD, Charnell Cromer, RN, MSN, Ayanna Walters, RN, BSN | Detroit, MI |
| Weill Cornell Chelsea CRS / NIAID (UM1AI068619) | Kristen Marks, MS, MD, Grant Ellsworth, MD, MS, Caroline Greene, ANP-BC, Sarah Galloway, BA, Shashi Kapadia, MD, MS, Elliot DeHaan, MD | New York, NY |
| Willis-Knighton Health System / WKB Family Medicine Associates | Clint Wilson, MD, Jason Milligan, MD, Danielle Raley, MD, Joseph Bocchini, MD | Bossier City, LA |
| Womack Army Medical Center | Bruce McClenathan, MD, Mary Hussain, BS, Evelyn Lomasney, MD, Evelyn Hall, MMS, PA-C, Sherry Lamberth, PharmD | Fort Bragg, NC |
| WR Clinsearch | Mark McKenzie, MD, Teresa Deese, Christy Schmeck, Vickie Leathers, Christy Sweet | Chattanooga, TN |

* Funding of institutions by the National Institute of Allergy and Infectious Diseases (NIAID) and/or research support by the National Center for Advancing Translational Science (NCATS), as indicated. All other institutions were funded by Office of the Assistant Secretary for Preparedness and Response, Biomedical Advanced Research and Development Authority. The

content of this publication is solely the responsibility of the authors and does not necessarily represent the official views of the funding sources.
